# Comparative Effectiveness of BNT162b2 versus Ad26.COV2.S for the Prevention of COVID-19 among Dialysis Patients

**DOI:** 10.1101/2021.10.21.21265339

**Authors:** Steven M. Brunelli, Scott Sibbel, Steph Karpinski, Gilbert Marlowe, Adam G. Walker, Jeffrey Giullian, David Van Wyck, Tara Kelley, Rachael Lazar, Meredith L. Zywno, Jeffrey J. Connaire, Amy Young, Francesca Tentori

**Affiliations:** DaVita Clinical Research, Minneapolis, Minnesota, USA; DaVita Inc, Denver, Colorado, USA

## Abstract

**Background:** mRNA-based SARS-CoV-2 vaccines have been shown to be highly effective among dialysis patients. Because individual vaccines may be differentially available or acceptable to patients, it is important to understand comparative effectiveness of other vaccines, such those based on adeno-virus technologies.

**Methods:** This retrospective study compared the clinical effectiveness of Ad26.COV2.S (Janssen/Johnson and Johnson) to BNT162b2 among dialysis patients. Patients initiating BNT162b2 (Pfizer/BioNTech) were matched 1:1 to Ad26.COV2.S recipients based on age, race, US state of residence, calendar week of first vaccine receipt, and history of COVID-19. The primary outcome was the comparative rate of COVID-19 considered over 3 follow-up intervals: weeks 1-3, 4-6, and ≥ 7 post-vaccination. In a subset of consented Ad26.COV2.S patients, blood samples were collected ≥28 days after vaccination and anti-SARS-Cov-2 immunoglobulin G antibodies were measured.

**Results:** There were 2659 matched pairs of patients who received a first dose of each vaccine. During weeks 1-3, incidence rates were 1.13 vs 1.39 per 1000 patient-weeks (pt-wks) for BNT162b2 and Ad26.COV2.S recipients, respectively (incident rate difference [IRD]: 0.25; 95% CI: -0.90, 1.36). During weeks 4-6, incidence rates were 0.78 vs 0.39 per 1000 pt-wks for BNT162b2 and Ad26.COV2.S recipients, respectively (IRD: -0.39; 95% CI: -1.16, 0.38). After week 7, incidence rates were 1.29 vs 1.39 per 1000 pt-wks for BNT162b2 and Ad26.COV2.S recipients, respectively (IRD: 0.10; 95% CI: -0.35, 0.55). Results were similar when considering only patients without a history of COVID-19 and among matched pairs in which BNT162b2 recipients completed the 2-dose regimen. SARS-CoV-2 antibodies were detected in 59.4% (95% CI: 53.0%-65.5%) of Ad26.COV2.S patients.

**Conclusion:** In a large real-world cohort of dialysis patients, no difference was detected in the clinical effectiveness of BNT162b2 and Ad26.COV2.S, despite an inconsistent antibody response to the latter. These data support the use of either agent in ongoing vaccination efforts in this population.

## INTRODUCTION

The population of patients receiving maintenance dialysis for end-stage kidney disease is enriched for risk factors for—and has borne a disproportionate burden of—infection with SARS-CoV-2 and severe COVID-19 manifestation.^1-4^ As such, efforts to vaccinate these patients and understand the effectiveness of vaccinations in this cohort are critical. At present, 3 vaccines have been approved or granted emergency use authorization by the US Food and Drug Administration: the mRNA-based BNT162b2 (Pfizer/BioNTech) and mRNA-1273 (Moderna) and the adenovirus-based Ad26.COV2.S (Janssen/Johnson & Johnson). To date, there are no comparative effectiveness data for these vaccines on clinical outcomes.

Dialysis providers enrolled as vaccine providers to increase access to SARS-CoV-2 vaccines;^5^ vaccines were received via an allocation from the US federal government (BNT162b2) as well as from state governments (any vaccine, including of Ad26.COV2.S). Typically, at any point of care, individual patients had access to only one vaccine (i.e., providers could not route a particular vaccine to a particular patient based on perceived need or patient preference). Because of the parallel nature of the federal and state allocations, at points in time patients living in a similar geographic area received different vaccines (i.e., BNT162b2 and Ad26.COV2.S), creating a natural experiment.

Overall vaccine uptake in the US dialysis population has been robust, with the majority of patients having received BNT162b2 or mRNA-1273, both of which have been demonstrated to be highly effective in preventing COVID-19.^6^ As vaccination efforts proceed, a meaningful number of dialysis patients have indicated hesitancy to receive either of these vaccines because they are mRNA-based; however, such patients do express a willingness to receive the adenovirus-based Ad26.COV2.S. Recent studies have demonstrated a lackluster serologic response to Ad26.COV2.S among dialysis patients, calling into question its effectiveness in this population.^7^ Thus, it is important to understand the effectiveness of Ad26.COV2.S to inform both ongoing vaccination efforts as well as clinical decisions for the care of dialysis patients who have already received this vaccine.

Therefore, we leveraged the natural experiment established by vaccine allocation programs to test the real-world comparative effectiveness of the adenovirus-based Ad26.COV2.S versus one of the mRNA-based vaccines (BNT162b2) in preventing clinical cases of COVID-19 among a large, representative cohort of US dialysis patients. Given that real world effectiveness is impacted by patient adherence (ie, to the 2^nd^ of two-dose regimen), we began by considering all matched BNT162b2 : AD26.COV2.S pairs, regardless of whether the latter followed through with a second dose. Recognizing that there is interest in effectiveness when regimens are followed as intended, we performed restricted analyses in which matched pairs were considered only if the BNT162b2 patient completed a 2-dose regimen (“completer pairs”). Furthermore, for each comparison, we performed separate analyses including and not including patients with a prior history of COVID-19.

## METHODS

### Comparative Effectiveness Analyses

This retrospective study was designed to compare 2 potential vaccination regimens: “use BNT162b2” versus “use Ad26.COV2.S.” BNT162b2 was chosen as the comparator messenger RNA vaccine (as opposed to mRNA-1273) because it had better geo-temporal overlap with AD26.COV2.S in our population. The study included dialysis patients who were ≥18 years and who received a first dose of BNT162b2 or a dose of Ad26.COV2.S between 01 January and 18 May 2021; the date of this dose was set as the index date. During the study period, approximately one-third of patient vaccinations occurred in dialysis facilities. For those patients who did not receive a vaccine in-clinic, vaccine information was ascertained through verbal attestation and review of vaccine cards. To account for geo-temporal variability in the intensity of the epidemic, BNT162b2 patients were exactly matched to Ad26.COV2.S patients on US state of residence and calendar week of vaccine receipt. Furthermore, to promote balance on susceptibility factors, BNT162b2 and Ad26.COV2.S patients were also matched on age (± 5 years), race, and history of prior COVID-19. Covariate balance between matched patients was assessed using descriptive statistics (means, SDs, counts, and proportions) and quantified in terms of standardized differences; standardized differences exceeding ± 10% indicate substantial imbalance.^8^

The primary outcome was a polymerase chain reaction (PCR)–confirmed clinical diagnosis of COVID-19. Clinical surveillance protocols at the dialysis organization have been described elsewhere.^9, 10^ To summarize, at the time of each clinic visit, all patients undergo a standardized COVID-19 screening. Patients who screen positive for COVID-19 symptoms, or who indicate a recent contact with COVID-19–infected individuals, immediately receive PCR testing. Additional surveillance procedures are in place to identify COVID-19 diagnoses made at other sites of care (e.g., emergency departments, hospitals) and to confirm and document PCR test results from these sites.

Patients were considered at-risk beginning on the day after index date, and remained at-risk until the earliest of documented COVID-19 diagnosis, death, loss to follow-up, or on study end (28 September 2021). The follow-up period was separated into 3 intervals based on a priori assumptions: weeks 1-3, 4-6, and ≥7, corresponding to the interval between doses, the period after the second dose and before development of immunity, and the period of presumed immunity for BNT162b2 recipients, respectively. Corresponding follow-up time intervals were used for Ad26.COV2.S. Comparative effectiveness was estimated using Poisson regression and expressed as incidence rate differences (IRD) and 95% CIs for each of the 3 follow-up periods. As standard, IRDs for which 95% CIs exclude zero were considered statistically significant. In addition, because finite statistical power increases the likelihood of type 2 errors, we further consider a “worst case” scenario for Ad26.COV2.S based on the upper confidence bound, which is a conservative approach since statistical imprecision implicitly “counts against” Ad26.COV2.S under this approach.

Separate analyses were performed in parallel among 4 nested populations: 1) all matched BNT162b2 pairs : Ad26.COV2.S; 2) matched pairs excluding those with preceding history of COVID-19; 3) matched pairs in whom BNT162b2 patients received a 2^nd^ dose (completer pairs); 4) completer pairs excluding those with preceding history of COVID-19. To avoid biases that would otherwise result from conditioning on future information, analyses of completer pairs considered only the follow-up period ≥ 7 weeks.

According to 45 Code of Federal Regulations part 46 from the US Department of Health and Human Services, this study was deemed exempt from institutional review board (IRB) or ethics committee approval. We adhered to the Declaration of Helsinki, and informed consent was not required. Data were derived from electronic health records. Socioeconomic indicators were derived from the 2020 US census and linked by zip code.^11^ All analyses were performed using Stata 10.0/MP (College Station, TX).

### Antibody Analysis

Elsewhere, we have reported high rates of seroresponse (98.1%) in dialysis patients completing the two dose regimen of BNT162b2.^6^ To estimate vaccine antibody response to Ad26.COV2.S, we conducted a prospective study among a subset of patients who received Ad26.COV2.S between 15 January and 25 March 2021. This study was reviewed and approved by Advarra IRB (Protocol # DCR 21-S-0016-00; approved on 19 March 2021). After patients provided written informed consent, a blood sample was collected 28 to 56 days after vaccine receipt to measure anti-SARS-CoV-2 antibodies. Blood samples were collected prior to a dialysis treatment in a 5 mL serum separation tube, clotted for 30 minutes, centrifuged, and refrigerated prior to shipment. All samples were processed at a centralized, accredited lab (DaVita Labs). Immunoglobulin G (IgG) was measured using an indirect chemiluminescence immunoassay for anti-SARS-CoV-2 IgG antibodies (Diazyme Laboratories, Inc), which detects antibodies against the SARS-CoV-2 spike and nucleocapsid proteins. Per the manufacturer’s recommendation, samples were scored IgG positive if the corresponding test reading was >1 arbitrary unit/mL, and negative otherwise. We previously used this assay for research purposes due to its selectivity for SARS-CoV-2 antibodies and low-levels of cross-reactivity to other coronaviruses or influenza viruses.^9, 10^ IgM levels were not measured based on pilot data demonstrating that measured IgM response was implausibly low in this population (<20% positive even in the 28-day period following documented SARS-CoV-2 infection).

## RESULTS

### Comparative Effectiveness among All Patients

We identified 2659 matched pairs of BNT162b2 and Ad26.COV2.S patients. Characteristics of patients in the matched sample were well-balanced (Table 1). Among BNT162b2 patients with at least 21 days of follow-up, 95.5% received a second BNT162b2 dose.

**Table 1.** Baseline characteristics of matched BNT162b2 and Ad26.COV2.S patients in the primary analyses ^a^

|  | All Patients |  |  | COVID-19 Naïve Patients |  |  |
| --- | --- | --- | --- | --- | --- | --- |
|  | BNT162b2<br>N = 2659 | Ad26.COV2.S<br>N = 2659 | Std Diff<sup>b</sup> | BNT162b2<br>N = 2465 | Ad26.COV2.S<br>N = 2465 | Std Diff<sup>b</sup> |
| **Age, years, mean ± SD** | 62.0 ± 12.2 | 62.1 ± 12.6 | +0.8% | 62.1 ± 12.3 | 62.2 ± 12.6 | +0.8% |
| **Female sex, %** | 43.1 | 41.5 | -3.2% | 43.1 | 41.4 | -3.4% |
| **Race, %** |  |  |  |  |  |  |
| White | 34.6 | 34.7 | +0.2% | 35.5 | 35.5 | +0.0% |
| Black | 32.8 | 32.8 | +0.0% | 33.3 | 33.3 | +0.0% |
| Hispanic | 20.6 | 20.7 | +0.2% | 18.7 | 18.9 | +0.5% |
| Asian | 3.6 | 3.7 | +0.5% | 3.8 | 3.9 | +0.5% |
| Other/unknown | 8.4 | 8.1 | -1.1% | 8.7 | 8.4 | -1.1% |
| **Dialysis modality, %** |  |  |  |  |  |  |
| In-center hemodialysis | 84.5 | 85.5 | +2.8% | 83.9 | 85.2 | +3.6% |
| Peritoneal dialysis | 13.5 | 12.9 | -1.8% | 14.0 | 13.1 | -2.6% |
| Home hemodialysis | 2.0 | 1.6 | -3.0% | 2.1 | 1.7 | -2.9% |
| **Prior history of COVID-19, %** | 7.3 | 7.3 | +0.0% | 0.0 | 0.0 | +0.0% |
| **Socioeconomic indicators <sup>c</sup>, %, mean ± SD** |  |  |  |  |  |  |
| Household poverty | 15.7 ± 9.2 | 15.2 ± 8.2 | -5.7% | 15.6 ± 9.3 | 15.1 ± 8.2 | -5.7% |
| Housing vacancy | 9.7 ± 6.6 | 10.4 ± 6.8 | +10.4% | 9.8 ± 6.6 | 10.4 ± 6.9 | +8.9% |
| 18-24 year olds without a high school diploma | 14.0 ± 8.6 | 14.0 ± 7.3 | +0.0% | 13.9 ± 8.6 | 14.0 ± 7.3 | +1.3% |
| Unemployment | 6.5 ± 3.8 | 6.2 ± 3.0 | -8.8% | 6.5 ± 3.8 | 6.2 ± 3.0 | -8.8% |

<sup>a</sup> US state of residence and index week matched exactly (not shown).<sup>b</sup> Std Diff, standardized difference. Standardized differences exceeding ± 10% indicate substantial imbalance.<sup>15</sup><sup>c</sup> Based on census data linked through zip code.

COVID-19 incidence rates over time after each vaccine administration are shown in Table 2. During weeks 1-3 of follow-up, incidence rates in the BNT162b2 and Ad26.COV2.S groups were 1.13 vs 1.39 diagnoses per 1000 pt-wks, respectively. During weeks 4-6 of follow-up, incidence rates in the BNT162b2 and Ad26.COV2.S groups were 0.78 vs 0.39 diagnoses per 1000 pt-wks, respectively. During weeks 7 and beyond, incidence rates in the BNT162b2 and Ad26.COV2.S groups were 1.29 vs 1.39 diagnoses per 1000 pt-wks, respectively. Figure 1A shows IRDs and 95% CIs during the 3 study follow-up periods. There were no statistically significant differences among IRDs during any of the follow-up periods. Based on the confidence limits, with 95% certainty, the number of additional COVID-19 diagnoses per week for each 1000 patients adopting a “use Ad26.COV2.S” (vs a “use BNT162b2”) would not be expected to exceed 1.36 in the first 3 weeks, 0.38 during weeks 4-6, and 0.55 beyond 7 weeks.

**Table 2.** COVID-19 diagnoses during follow-up for patients treated with BNT162b2 or Ad26.COV2.S considering all matched pairs

|  | Weeks 1-3 |  | Weeks 4-6 |  | Weeks 7+ |  |
| --- | --- | --- | --- | --- | --- | --- |
|  | BNT162b2 | Ad26.COV2.S | BNT162b2 | Ad26.COV2.S | BNT162b2 | Ad26.COV2.S |
| **All Patients** |  |  |  |  |  |  |
| **Patients** | 2659 | 2659 | 2574 | 2574 | 2492 | 2492 |
| **At-risk time (pt-wks)** | 7942 | 7934 | 7679 | 7672 | 50,245 | 50,240 |
| **COVID-19 diagnoses** | 9 | 11 | 6 | 3 | 65 | 70 |
| **Incidence rate (per 1000 pt-wks)** | 1.13 | 1.39 | 0.78 | 0.39 | 1.29 | 1.39 |
| **COVID-19 Naïve Patients** |  |  |  |  |  |  |
| **Patients** | 2465 | 2465 | 2385 | 2385 | 2310 | 2310 |
| **At-risk time (pt-wks)** | 7364 | 7354 | 7117 | 7108 | 46,551 | 46,572 |
| **COVID-19 diagnoses** | 9 | 10 | 6 | 3 | 65 | 70 |
| **Incidence rate (per 1000 pt-wks)** | 1.22 | 1.36 | 0.84 | 0.42 | 1.40 | 1.50 |

**Figure 1.**
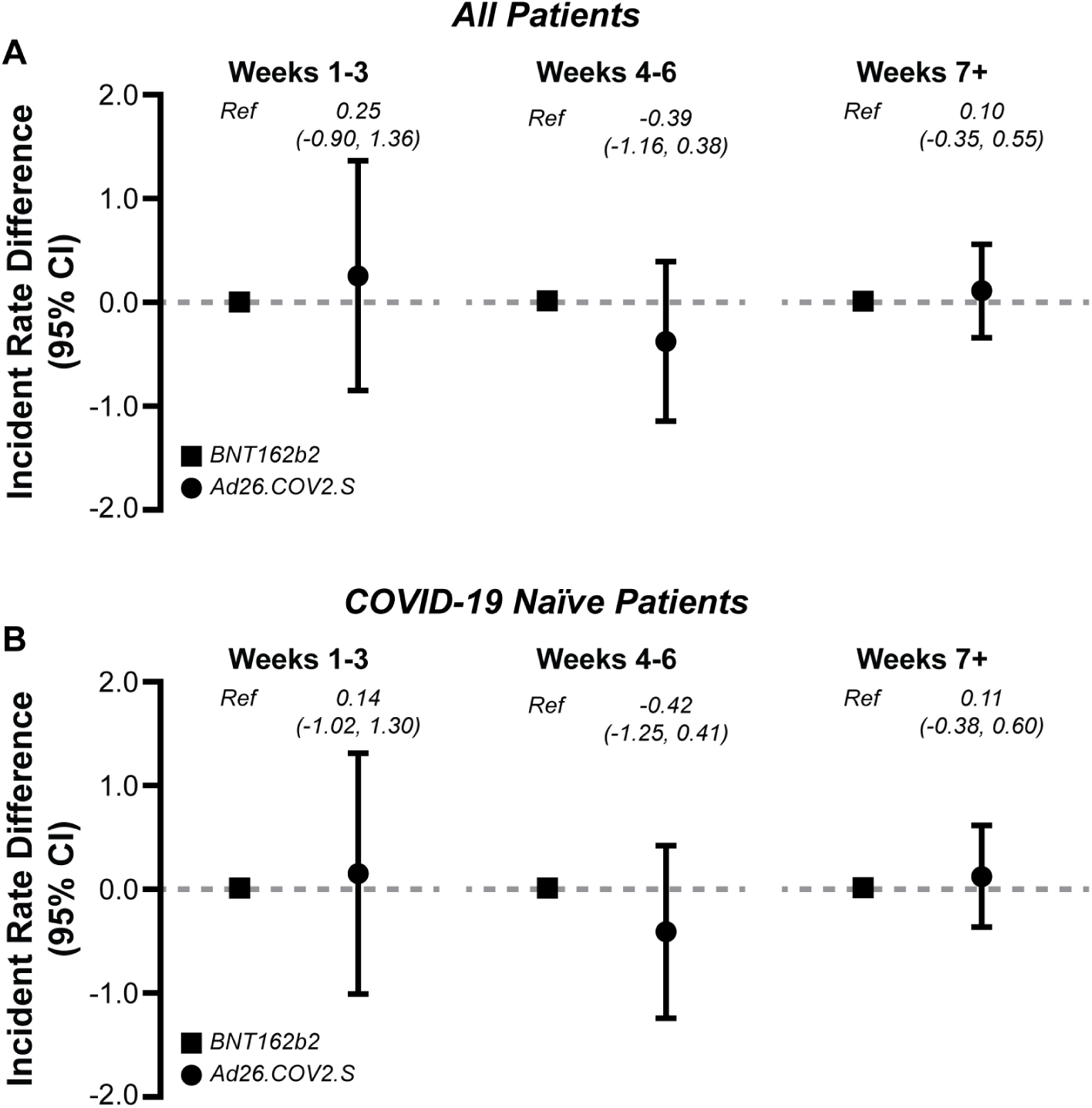
Comparative effectiveness of BNT162b2 and Ad26.COV2.S during follow-up among (A) all matched pairs and. (B) the subset of matched pairs with no prior history of COVID-19.

### Comparative Effectiveness among Patients with No Prior COVID-19 History

When we excluded pairs with a prior history of COVID-19, there remained 2465 matched pairs of BNT162b2 and Ad26.COV2.S patients. Characteristics were similar to those seen in the parent cohort and were well-balanced across groups (Table 1). COVID-19 incidence rates by vaccine type over follow-up time are shown in Table 2. During weeks 1-3 of follow-up, incidence rates in the BNT162b2 and Ad26.COV2.S groups were 1.22 vs 1.36 diagnoses per 1000 pt-wks, respectively. During weeks 4-6 of follow-up, incidence rates in the BNT162b2 and Ad26.COV2.S groups were 0.84 vs 0.42 diagnoses per 1000 pt-wks, respectively. During weeks 7 and beyond, incidence rates in the BNT162b2 and Ad26.COV2.S groups were 1.40 vs 1.50 diagnoses per 1000 pt-wks, respectively. Figure 1B shows IRDs and 95% CIs during the 3 study follow-up periods for patients who did not have a history of COVID-19. There were no statistically significant differences among IRDs during any of the follow-up periods. Based on the confidence limits, with 95% certainty, the number of additional COVID-19 diagnoses generated per week for each 1000 patients without a prior COVID-19 history, adopting a “use Ad26.COV2.S” (vs a “use BNT162b2”) strategy would not be expected to exceed 1.30 in the first 3 weeks, 0.41 during weeks 4-6, and 0.60 beyond 7 weeks.

### Comparative Effectiveness among Vaccine Dose Regimen Completers

Infection rates by vaccine type when analyses were restricted to matched pairs in which BNT162b2 patients completed a 2-dose series are shown in Table 3. Incidence rates for follow-up weeks 7 and beyond were 1.23 vs 1.34 diagnoses per 1000 pt-wks in the BNT162b2 and Ad26COV2.S groups, respectively. This difference was not statistically significant: IRD (95% CI) 0.11 (−0.34, 0.56; *P*>0.05). Based on the confidence limits, with 95% certainty, the number of additional COVID-19 diagnoses generated per week for each 1000 patients adopting a “use Ad26.COV2.S” (vs a “use BNT162b2”) strategy would not be expected to exceed 0.56. When consideration was limited to the subset of completer pairs with no prior history of COVID-19, results were nearly identical (Table 3).

**Table 3.** COVID-19 diagnoses during follow-up weeks ≥ 7 for matched pairs in whom BNT162b2 patients completed the two-dose vaccine series

|  | All Patients |  | COVID-19 Naïve Patients |  |
| --- | --- | --- | --- | --- |
|  | BNT162b2 | Ad26.COV2.S | BNT162b2 | Ad26.COV2.S |
| **Patients** | 2404 | 2404 | 2230 | 2230 |
| **At-risk time (pt-wks)** | 48,748 | 48,518 | 45,180 | 45,013 |
| **COVID-19 diagnoses** | 60 | 65 | 60 | 65 |
| **Incidence rate (per 1000 pt-wks)** | 1.23 | 1.34 | 1.33 | 1.44 |
| **IRD (95% CI)** | Ref | 0.11 (-0.34, 0.56) | Ref | 0.12 (-0.37, 0.60) |

### Antibody Response to Ad26.COV2.S

We measured antibodies in 244 patients who received Ad26.COV2.S. Median and interquartile range of time from Ad26.COV2.S receipt to blood sampling was 36.5 [34, 41] days. IgG antibodies were detected in 59.4% (95% CI: 53.0%-65.5%) of patients who were vaccinated with Ad26.COV2.S.

## DISCUSSION

In this comparative effectiveness study in patients on maintenance dialysis, we observed no difference in clinical effectiveness between vaccination with Ad26.COV2.S versus BNT162b2. Results were similar when we only considered patients who did not have a prior history of COVID-19, in whom vaccine effectiveness should therefore be minimally impacted by naturally acquired immunity. These results contribute important information to guide clinical practice for dialysis patients, at a time when there is some speculation that Ad26.COV2.S may be substantially inferior to the mRNA SARS-CoV-2 vaccines.

In clinical trials, the efficacy of Ad26.COV2.S was estimated to be 67%, whereas the mRNA vaccines had reported efficacies of 94% to 95%.^12-14^ It is important to note that these trials did not include dialysis patients. Moreover, comparing clinical efficacy estimates across clinical trials may not be valid because of differences in patient populations included in each trial, background rates of SARS-CoV-2 transmission, and surveillance procedures. Finally, efficacy results from clinical trials may not accurately reflect effectiveness in the real-world setting due to patient selection and study oversight, the latter particularly of concern when an opportunity for differential adherence exists (i.e., the need for a second vaccination with mRNA vaccines). In the dialysis population, concerns about the effectiveness of Ad26.COV2.S are further compounded by recent evidence that antibody responses to Ad26.COV2.S are less robust and consistent than other SARS-CoV-2 vaccines.^7^ In the present study, we observed that approximately 60% of dialysis patients vaccinated with Ad26.COV2.S had detectable SARS-CoV-2 antibodies. This is comparatively lower than in our previous report in patients who received mRNA vaccines (96-98%).^6^ Obviously, the serologic results and clinical effectiveness for Ad26.COV2.S is discordant. The reason why the serologic data do not accurately reflect the clinical effectiveness of this vaccine remain unknown.

Given these uncertainties, it has been unclear how best to approach vaccination among dialysis patients who, either by circumstance (i.e., vaccine availability) cannot—or, by personal choice, will not—receive an mRNA-based vaccine. Moreover, the appropriate course of action for patients who have received Ad26.COV2.S remains to be determined. In this regard, our results should be seen as reassuring. In fact, using the upper 95% confidence bound (which yields a more conservative estimate due to statistical imprecision), our estimates suggest that the excess number of COVID-19 diagnoses following a “use Ad26.COV2.S” (versus “use BNT162b2”) approach should not exceed 0.55 per week per 1000 patients treated from week 7 on (i.e., the time period in which both vaccines should be at maximal effectiveness). In addition, because the period of time between first vaccination and presumed immunity is consequential both for patients and logistics of care delivery, we also provide estimates for earlier time periods; these are likewise reassuring in that Ad26.COV2.S appeared equivalent or (for weeks 4-6) trended toward superior, albeit the latter was not statistically significant due to a low number of events. Our primary analysis included all patients who began a vaccine regimen, regardless of whether individual BNT162b2 patients completed the 2-dose series; this deliberate choice was made to reflect the reality that some patients do not follow through with a second dose, with real-world consequences on effectiveness. Nevertheless, follow-through rates were very high, and Ad26.COV2.S remained noninferior, even in sensitivity analyses restricted to patients who received both BNT162b2 doses.

The strengths of this study include a large sample size, a representative patient population, and a robust statistical matching procedure to account for the geo-temporal nature of the COVID-19 pandemic and vaccine rollout in the United States. The study should be viewed in light of the following limitations. As with any observational study, there is the possibility of residual confounding and bias (e.g., misclassification of exposure status for patients reporting vaccinations outside of the clinic). Data limitations precluded the ability to study COVID-19-related hospitalizations as we well as severity of illness and asymptomatic infections, or to study side effects of vaccines. Low case counts also precluded study of case fatality. Given the fluctuating nature of background COVID-19 rates in the US population over the course of follow-up, COVID-19 rates within an exposure group should not be compared across follow-up periods, but rather limited to within period comparisons across exposure groups. Finally, we could not compare the effectiveness of these vaccines against specific SARS-CoV-2 variants.

In summary, our results demonstrate equivalent effectiveness of Ad26.COV2.S to an mRNA-based vaccine (BNT162b2) in dialysis patients. These results support the continued use of Ad26.COV2.S as a SARS-CoV-2 vaccine option in this vulnerable population.

## Data Availability

All data produced in the present study are proprietary and property of a HIPAA covered entity and therefore cannot be shared

## AUTHOR CONTRIBUTIONS

S.M. Brunelli, S. Sibbel, S. Karpinski, J. Guillian, and D. Van Wyck designed the study. Clinical/logistical leadership and oversight provided by T. Kelley, J.J. Connaire, F. Tentori, A. Young., and S.M. Brunelli. G. Marlowe, R. Lazar, and M. L. Zywno prepared the analytic data files. S. M. Brunelli performed the analysis. S.M. Brunelli, S. Sibbel, S. Karpinski, A.G. Walker, J. Guillian, D. Van Wyck, and F. Tentori interpreted the findings. S.M. Brunelli, A.G. Walker, and F. Tentori drafted the manuscript. The manuscript was reviewed and approved by all authors prior to submission.

## ACKNOWLEDGMENTS

The authors would like to thank the patients and staff of DaVita, Inc, without whom this research would not have been possible.

